# ADOLESCENT GIRLS’ KNOWLEDGE, ATTITUDES, AND PREVENTIVE PRACTICES TOWARD HIV: EVIDENCE FROM A SCHOOL-BASED SURVEY

**DOI:** 10.64898/2026.09.15.26363184

**Authors:** Stella I Ohaike, Rita C Emeh, Adaeze I Onyekwelu, Kadiree E Fatai

## Abstract

**Background:** Adolescent girls remain a priority population in HIV prevention efforts in sub-Saharan Africa due to heightened biological, social, and structural vulnerabilities. In Nigeria, schools serve as critical platforms for HIV education; however, evidence suggests that awareness alone may not be sufficient to ensure consistent preventive practices among adolescents. Understanding the interplay between knowledge, attitudes, and behaviors is essential for designing effective interventions. This study examined HIV-related knowledge, attitudes, and preventive practices among female secondary school students in an urban Nigerian setting.

**Methods:** A cross-sectional, school-based survey was conducted among 150 female adolescents aged 14-18 years attending a public girls’ secondary school in southeastern Nigeria. Participants were recruited using a convenience sampling approach. Data were collected using a structured questionnaire that assessed HIV transmission knowledge, common misconceptions, attitudes toward prevention and testing, perceived self-efficacy, and reported preventive practices. The instrument was pilot-tested for internal consistency prior to data collection. Descriptive statistics were used for analysis. Adequate HIV knowledge was defined as achieving at least 70% correct responses across knowledge items.

**Results:** Overall, 72.4% of respondents demonstrated adequate knowledge of HIV. Correct identification of sexual transmission was high (90.0%), as was awareness of blood-related transmission (85.3%). However, notable misconceptions persisted: 27.3% believed HIV could be transmitted through casual contact, and 21.9% perceived HIV as a condition affecting only specific groups. Attitudes toward HIV prevention were moderately positive, with 68.0% expressing willingness to undergo HIV testing and 61.3% reporting confidence in negotiating safer behaviours. Preventive practices were less robust; although 74.6% acknowledged the effectiveness of condoms, only 58.7% accurately identified all routes of mother-to-child transmission. Information sources were predominantly school-based (79.3%), followed by social media (65.4%) and parents or guardians (49.8%).

**Conclusions:** The findings reveal moderate HIV knowledge and generally positive attitudes among adolescent girls, alongside persistent misconceptions and gaps in preventive practices. Strengthening school-based HIV prevention efforts should prioritize correcting misinformation, enhancing practical prevention skills, and leveraging both formal education and digital platforms to support informed and sustained protective behaviors among adolescents.

## Introduction

Despite substantial advances in HIV prevention, diagnosis, and antiretroviral therapy, HIV remains a major global public health challenge. Globally, an estimated 39.9 million people were living with HIV in 2023, with approximately 1.3 million new infections recorded during the year, highlighting the persistent burden of the epidemic despite significant progress in HIV control (Joint United Nations Programme on HIV/AIDS [UNAIDS], 2024). Adolescents and young people continue to experience disproportionate HIV-related morbidity and mortality due to limited access to age-appropriate prevention, testing, and treatment services (Dzinamarira & Moyo, 2024). Among this population, adolescent girls and young women (AGYW) remain particularly vulnerable. In 2023, approximately 210,000 AGYW aged 15–24 years acquired HIV globally, representing about 4,000 new infections weekly, with the majority occurring in sub-Saharan Africa (UNAIDS, 2024). Although progress has been made toward global HIV targets, current trends remain insufficient to achieve the goal of ending AIDS as a public health threat by 2030.

Sub-Saharan Africa remains the epicentre of the global HIV epidemic and accounts for the majority of new HIV infections among AGYW. Nearly four out of every five new infections in this population occur within the region (UNAIDS, 2024). The heightened vulnerability of AGYW reflects a complex interaction of biological, social, and structural factors. Biological susceptibility, including increased vulnerability to sexually transmitted infections, combines with gender inequality, poverty, age-disparate relationships, gender-based violence, and limited autonomy to increase HIV risk and reduce the ability of girls to negotiate safer sexual practices (Murewanhema et al., 2022). These realities highlight the need for integrated HIV prevention strategies that address both biomedical and social determinants of HIV.

Nigeria remains central to the HIV response in sub-Saharan Africa because of its large population and substantial HIV burden. Approximately 1.9 million Nigerians are living with HIV, making the country one of the highest-burden settings globally (National Agency for the Control of AIDS [NACA], 2019). Adolescents, particularly girls, remain vulnerable because of socioeconomic disadvantage, gender inequality, limited access to youth-friendly sexual and reproductive health services, and persistent HIV-related stigma. In response, Nigeria has implemented adolescent-focused interventions such as the National HIV and AIDS Strategic Framework, the National Policy on HIV Prevention, Family Life and HIV Education (FLHE), and comprehensive sexuality education programmes aimed at improving HIV awareness and reducing risky sexual behaviours. However, persistent misconceptions, stigma, low HIV testing uptake, and inadequate comprehensive HIV knowledge continue to undermine prevention efforts (Akande et al., 2024; Badru et al., 2020).

Understanding these challenges requires examining not only awareness but also behavioural determinants of HIV prevention. Knowledge, attitudes, and practices (KAP) provide a useful framework for understanding health-related behaviours and designing public health interventions. The KAP framework suggests that knowledge may foster positive attitudes and encourage protective behaviours, although behaviour is also influenced by self-efficacy, perceived risk, social norms, and environmental factors (Launiala, 2009). In the context of HIV prevention, comprehensive knowledge is essential for informed decision-making, yet knowledge alone does not consistently translate into safer sexual behaviour or increased uptake of HIV prevention services (Badejo et al., 2024). Misconceptions and misinformation may further reinforce stigma, reduce perceived susceptibility, and weaken preventive behaviour (Abate et al., 2020; Adekunjo et al., 2021).

Evidence from sub-Saharan Africa indicates that HIV awareness among adolescents has improved following sustained school-based and community-based education programmes. Nevertheless, comprehensive HIV knowledge remains inconsistent, while misconceptions, poor condom negotiation skills, low self-efficacy, and stigma continue to limit behavioural adoption of preventive strategies (Abate et al., 2020; Badejo et al., 2024). These findings suggest that awareness alone is insufficient to produce meaningful behavioural change.

Despite growing literature, important evidence gaps remain. Most Nigerian studies have focused on mixed-sex adolescent populations or isolated dimensions of HIV knowledge, with limited attention to the interrelationships among knowledge, misconceptions, attitudes, self-efficacy, and preventive practices among female secondary school students. Furthermore, few comprehensive school-based studies have been conducted in southeastern Nigeria, where sociocultural and educational factors may uniquely influence HIV-related behaviours. Addressing these gaps is essential for developing gender-responsive, evidence-informed HIV prevention strategies. Therefore, this study assessed HIV-related knowledge, attitudes, misconceptions, and preventive practices among female secondary school students in southeastern Nigeria to generate evidence for school-based HIV prevention interventions and adolescent health programmes.

### Methods Study Design

This study employed a descriptive cross-sectional design using a school-based survey to assess HIV-related knowledge, attitudes, misconceptions, and preventive practices among adolescent girls. A cross-sectional design was considered appropriate because it enables the assessment of health-related knowledge, attitudes, and behaviours within a defined population at a single point in time without manipulation of the study environment, making it suitable for estimating prevalence and describing health-related outcomes (Setia, 2016). This study was reported in accordance with the Strengthening the Reporting of Observational Studies in Epidemiology (STROBE) guideline for cross-sectional studies (von Elm et al., 2007).

### Study Setting

The study was conducted in December 2025 in Enugu South Local Government Area, located in Enugu State, southeastern Nigeria. Enugu State is one of the five states in southeastern Nigeria and comprises both urban and rural communities with an established network of public and private secondary schools under the supervision of the Enugu State Ministry of Education.

Enugu South Local Government Area is predominantly urban and serves as one of the major educational and administrative centres of the state. The survey was conducted in a public girls’ secondary school within the Local Government Area. The school provides secondary education to adolescent girls from diverse socioeconomic backgrounds and implements the national secondary school curriculum, including the Family Life and HIV Education (FLHE) programme, which incorporates age-appropriate instruction on HIV prevention, reproductive health, and healthy lifestyle practices (Federal Ministry of Education Nigeria & National Educational Research and Development Council, 2003).

### Study Population

The study population comprised female adolescents aged 14–18 years enrolled in the selected public secondary school in Enugu South Local Government Area. Adolescence is a critical developmental stage characterized by biological, psychological, and social changes that may increase vulnerability to HIV infection, particularly among adolescent girls in sub-Saharan Africa (World Health Organization, 2024; Joint United Nations Programme on HIV/AIDS, 2024).

Eligible participants were female students within the specified age range who were present in school during the study period and willing to participate. Students who were absent during data collection, declined participation, or did not obtain parental consent or provide assent were excluded from the study.

### Sampling Technique

A convenience sampling technique was employed to recruit study participants. Following approval from the school authorities, eligible students who met the inclusion criteria were invited to participate during scheduled school hours. The objectives, procedures, potential benefits, and voluntary nature of the study were explained to all eligible students before recruitment.

Students who provided written assent and whose parents or guardians provided written informed consent were enrolled consecutively until the target sample size of 150 participants was achieved.

### Data Collection Instrument

Data were collected using a structured, self-administered questionnaire developed for this study and adapted from validated adolescent HIV knowledge and behavioural assessment instruments to suit the Nigerian secondary school context (Folasayo et al., 2017; Badru et al., 2020). The questionnaire consisted of 35 items organized into six sections.

The first section collected participants’ sociodemographic characteristics, including age and class level. The second section assessed HIV knowledge using 10 items covering HIV transmission, prevention, treatment, and mother-to-child transmission. Each item was answered as *True*, *False*, or *Don’t know*, with one point awarded for each correct response and zero points assigned to incorrect or “Don’t know” responses.

The third section assessed attitudes toward HIV prevention and people living with HIV using six Likert-scale items ranging from *Strongly disagree* to *Strongly agree*. The fourth section evaluated perceived risk, previous HIV education, self-efficacy, willingness to undergo HIV testing, and HIV-related behavioural intentions using six structured items. The fifth section assessed participants’ sources of HIV information using multiple-response items, while the sixth section assessed preventive health behaviours, counselling readiness, and linkage-to-care intentions using Likert-scale items.

Knowledge scores were summed and converted into percentages. Participants who achieved at least 70% correct responses across the knowledge items were classified as having adequate HIV knowledge, whereas those scoring below 70% were classified as having inadequate HIV knowledge.

### Validity and Reliability

The questionnaire underwent face and content validation prior to data collection. The draft instrument was reviewed by experts in public health and adolescent health to assess the clarity, relevance, and appropriateness of the questionnaire items. Their recommendations informed minor revisions to improve item wording and overall content suitability.

The revised questionnaire was subsequently pilot-tested among adolescents attending a secondary school not included in the main study to evaluate clarity, comprehensibility, and feasibility of administration. Feedback obtained during the pilot study informed the final version of the questionnaire used for data collection. Internal consistency reliability was assessed using Cronbach’s alpha, which yielded a coefficient of 0.82, indicating good reliability.

### Outcome Measures

The primary outcome measure was HIV-related knowledge. Adequate HIV knowledge was defined as achieving at least 70% correct responses across the knowledge assessment items, while scores below 70% were classified as inadequate HIV knowledge.

Secondary outcome measures included attitudes toward HIV prevention and people living with HIV, perceived self-efficacy, preventive health behaviours, common misconceptions regarding HIV transmission, and sources of HIV information. Misconceptions were assessed based on incorrect beliefs regarding HIV transmission, including casual contact and mosquito bites. Attitudes and self-efficacy were measured using Likert-scale responses, with higher scores indicating more positive attitudes and greater confidence in adopting preventive behaviours.

### Data Collection Procedure

Data collection was conducted during scheduled school hours following approval from the school authorities. Eligible participants were informed about the purpose and procedures of the study before administration of the questionnaire. Students who provided written assent and whose parents or guardians had provided written informed consent were enrolled in the study.

The questionnaire was administered in a classroom setting under the supervision of the investigators to ensure standardization and provide clarification where necessary. Participants completed the questionnaires anonymously using unique identification numbers rather than names to maintain confidentiality. Completed questionnaires were checked for completeness immediately after collection before secure storage for data entry and analysis.

### Statistical Analysis

Data were entered, cleaned, and analysed using IBM SPSS Statistics version 26.0. Descriptive statistical methods were used to summarize the data. Categorical variables were presented as frequencies and percentages, while continuous variables were summarized using means and standard deviations where appropriate.

Knowledge scores were computed by summing correct responses to the HIV knowledge items and converting the total scores into percentages. Participants with knowledge scores of ≥70% were classified as having adequate HIV knowledge, whereas those scoring below 70% were classified as having inadequate HIV knowledge. The analysed data were presented in tables and descriptive summaries.

### Ethical Considerations

Ethical approval for this study was obtained from the University of Nigeria Teaching Hospital Research Ethics Committee, Ituku-Ozalla, Nigeria (Approval No.: [Insert Approval Number]) prior to data collection. Permission to conduct the study was also obtained from the Enugu State Ministry of Education and the management of the participating school.

Written informed consent was obtained from parents or guardians, while written assent was obtained from all participants before enrolment in the study. Participation was voluntary, and participants were informed of their right to withdraw from the study at any time without any consequences. Confidentiality was maintained through the use of anonymous questionnaires and unique identification codes. The study was conducted in accordance with the ethical principles of the Declaration of Helsinki.

## Results

### Sociodemographic Characteristics of Respondents

A total of 150 female adolescents participated in the study, representing a response rate of 100%. The respondents were aged between 14 and 18 years, with a mean age of 16.2 ± 1.3 years, indicating that the study population largely consisted of mid-to-late adolescents.

Age distribution showed that respondents aged 16–17 years constituted the largest proportion (60; 40.0%), suggesting that a substantial number of participants were within the age bracket commonly associated with increased vulnerability to sexual health risks. Participants aged 14–15 years accounted for 52 (34.7%), while those aged 18 years represented 38 (25.3%) of the study population.

Regarding educational level, more than half of the respondents (82; 54.7%) were enrolled in senior secondary classes, while 68 (45.3%) were in junior secondary classes. This relatively balanced distribution allowed representation across early and late secondary school years.

In terms of religious affiliation, the majority of respondents identified as Christians (118; 78.7%), whereas 32 (21.3%) identified as Muslims, reflecting the predominant religious demographics of the study setting.

Assessment of household structure revealed that 94 respondents (62.7%) lived with both parents, indicating relatively stable family support structures for most participants. Meanwhile, 34 (22.7%) lived with a single parent, and 22 (14.6%) resided with guardians or relatives.

Parental educational attainment showed that 87 respondents (58.0%) reported that at least one parent had completed tertiary education, while 46 (30.7%) reported parental secondary education and 17 (11.3%) reported primary education or below. This suggests that a considerable proportion of respondents came from relatively educated households, which may influence access to health-related information.

**Table 1.** Sociodemographic Characteristics of Respondents.

| Variable | Frequency | Percentage |
| --- | --- | --- |
| 14–15 years | 52 | 34.7 |
| 16–17 years | 60 | 40.0 |
| 18 years | 38 | 25.3 |
| Junior secondary | 68 | 45.3 |
| Senior secondary | 82 | 54.7 |
| Christian | 118 | 78.7 |
| Muslim | 32 | 21.3 |
| Both parents | 94 | 62.7 |
| Single parent | 34 | 22.7 |
| Guardian/relative | 22 | 14.6 |

### HIV Knowledge Among Respondents

Assessment of HIV-related knowledge showed that respondents generally demonstrated a satisfactory level of awareness regarding HIV transmission, prevention, and management. Overall, 108 respondents (72.0%) achieved the predefined threshold for adequate HIV knowledge (≥70% correct responses), indicating that nearly three-quarters possessed acceptable knowledge levels. However, 42 respondents (28.0%) had inadequate knowledge, suggesting that substantial knowledge gaps remained among a notable minority of participants.

Knowledge of the major routes of HIV transmission was particularly high. The vast majority of respondents (135; 90.0%) correctly identified unprotected sexual intercourse as a major route of HIV transmission. Similarly, 128 respondents (85.3%) recognized that HIV could be transmitted through contact with infected blood, including unsafe blood transfusion, sharing needles, and contaminated sharp objects.

Knowledge regarding HIV prevention strategies was also relatively high. A total of 121 respondents (80.7%) correctly identified abstinence from sexual activity as an effective preventive measure, while 112 respondents (74.7%) acknowledged that consistent condom use significantly reduces the risk of HIV transmission. These findings indicate that most respondents were aware of major preventive strategies.

In relation to clinical understanding of HIV, 117 respondents (78.0%) correctly stated that a healthy-looking person can still be infected with HIV, demonstrating moderate awareness that physical appearance alone cannot determine HIV status. Additionally, 104 respondents (69.3%) understood that although HIV currently has no complete cure, treatment can effectively control the infection and improve quality of life.

Knowledge regarding mother-to-child transmission (MTCT) was comparatively lower than other knowledge domains. Only 88 respondents (58.7%) correctly identified all routes of MTCT, including transmission during pregnancy, childbirth, and breastfeeding. This finding suggests weaker knowledge in more specific and less frequently emphasized aspects of HIV education.

Furthermore, 96 respondents (64.0%) recognized the importance of early HIV testing for prompt treatment and prevention of complications. Although this proportion reflects moderate awareness, it also suggests that more than one-third of respondents lacked adequate understanding of the benefits of early diagnosis.

Overall, the findings suggest relatively good awareness of common HIV transmission routes and prevention methods. However, knowledge of more complex or less frequently discussed areas particularly mother-to-child transmission and HIV treatment remained comparatively lower, indicating the need for more comprehensive HIV education.

**Table 2.** HIV Knowledge Among Respondents.

| Variable | Frequency (%) |
| --- | --- |
| Sexual intercourse can transmit HIV | 135 (90.0) |
| Blood exposure can transmit HIV | 128 (85.3) |
| Sharing needles/sharp objects transmits HIV | 124 (82.7) |
| Abstinence prevents HIV | 121 (80.7) |
| Healthy-looking person can have HIV | 117 (78.0) |
| Condom use reduces HIV risk | 112 (74.7) |
| HIV has no complete cure but is manageable | 104 (69.3) |
| Early HIV testing is beneficial | 96 (64.0) |
| Correct MTCT knowledge | 88 (58.7) |
| Adequate overall knowledge | 108 (72.0) |

### Misconceptions Regarding HIV Transmission

Despite the generally satisfactory level of HIV knowledge observed among respondents, several misconceptions regarding HIV transmission persisted. These misconceptions highlight important gaps in understanding that may contribute to stigma, discrimination, and inaccurate personal risk assessment.

A total of 41 respondents (27.3%) incorrectly believed that HIV could be transmitted through casual social contact, such as hugging, handshakes, or sharing utensils. This indicates that more than one-quarter of respondents still held inaccurate beliefs regarding non-transmissible forms of contact. Such misconceptions may contribute to fear-based attitudes and social distancing from persons living with HIV.

Similarly, 29 respondents (19.3%) believed that mosquito bites could transmit HIV. Although this misconception was less prevalent than casual-contact misconceptions, it nevertheless reflects persistent misinformation regarding biological transmission mechanisms.

In addition, 33 respondents (22.0%) perceived HIV as a disease affecting only specific high-risk groups, such as sex workers, people who inject drugs, or individuals perceived to engage in risky sexual behaviour. This belief reflects stereotypical thinking and may reduce perceived susceptibility among adolescents who do not identify with these groups, thereby undermining preventive behaviour.

Further misconceptions were also observed. 26 respondents (17.3%) believed that HIV could be transmitted through sharing toilets or bathrooms, while 24 respondents (16.0%) incorrectly thought that sharing food or drinks with an infected person could result in transmission. These misconceptions indicate continued confusion about everyday social interactions and HIV transmission risk.

Additionally, 18 respondents (12.0%) believed HIV could be caused by supernatural forces such as curses or witchcraft, suggesting that cultural myths and traditional beliefs may still influence understanding among some adolescents.

Overall, the findings demonstrate that although most respondents possessed adequate general knowledge about HIV, misconceptions remained evident across several domains. These persistent false beliefs underscore the need for school-based HIV education programmes to address myths directly and reinforce accurate understanding of HIV transmission.

**Table 3.** Misconceptions Regarding HIV Transmission.

| **Misconception** | **Frequency (%)** |
| --- | --- |
| Casual contact (hugging/handshake) transmits HIV | 41 (27.3) |
| HIV affects only specific groups | 33 (22.0) |
| Mosquito bites transmit HIV | 29 (19.3) |
| Sharing toilets transmits HIV | 26 (17.3) |
| Sharing food/drinks transmits HIV | 24 (16.0) |
| HIV caused by witchcraft/curses | 18 (12.0) |

Assessment of respondents’ attitudes toward HIV prevention and testing revealed generally moderately positive attitudes, although some stigma-related concerns remained evident.

A majority of respondents (102; 68.0%) expressed willingness to undergo HIV testing, indicating a relatively favorable disposition toward early diagnosis and health-seeking behaviour. This suggests that most participants recognized the importance of knowing one’s HIV status as part of preventive healthcare.

Similarly, 114 respondents (76.0%) agreed that regular HIV screening is important, reflecting broad awareness of HIV testing as a critical component of prevention and early intervention. In addition, 109 respondents (72.7%) believed that practicing preventive measures such as abstinence or condom use reflects responsible health behaviour.

Regarding perceived vulnerability, 87 respondents (58.0%) agreed that adolescents are personally at risk of HIV infection if preventive measures are not taken. This suggests moderate awareness of personal susceptibility, although a substantial proportion may still underestimate their individual risk.

Attitudes toward people living with HIV (PLWH) were generally positive but revealed residual stigma. 121 respondents (80.7%) reported willingness to study or interact in the same environment with persons living with HIV, indicating relatively good social acceptance. However, 29 respondents (19.3%) still reported discomfort interacting closely with PLWH, reflecting persistent discriminatory attitudes among a minority of respondents.

Stigma-related concerns remained evident in relation to diagnosis and disclosure. 36 respondents (24.0%) reported fear of stigma or discrimination if diagnosed with HIV, suggesting that social judgment remains a potential barrier to testing and disclosure. Furthermore, 43 respondents (28.7%) expressed reluctance to openly disclose a positive HIV status to peers or extended family members.

**Table 4.** Attitudes Toward HIV Prevention and Testing.

| Variable | Frequency (%) |
| --- | --- |
| Willing to undergo HIV testing | 102 (68.0) |

| **Variable** | **Frequency (%)** |
| --- | --- |
| Regular HIV testing is important | 114 (76.0) |
| Preventive behaviour is important | 109 (72.7) |
| Perceived personal susceptibility | 87 (58.0) |
| Comfortable interacting with PLWH | 121 (80.7) |
| Fear stigma after diagnosis | 36 (24.0) |
| Discomfort with PLWH | 29 (19.3) |
| Reluctance to disclose HIV status | 43 (28.7) |

Overall, the findings indicate that respondents demonstrated largely positive attitudes toward HIV prevention and testing. Nevertheless, concerns related to stigma, disclosure, and social acceptance remain important barriers that may influence testing uptake and preventive behaviour.

### Preventive Practices and Self-Efficacy

Assessment of preventive practices and self-efficacy revealed comparatively weaker performance than the knowledge and attitude domains. Although respondents generally demonstrated reasonable awareness of preventive strategies, this knowledge did not consistently translate into confidence or protective behavioural practices.

A total of 112 respondents (74.7%) acknowledged that consistent condom use reduces the risk of HIV transmission, indicating that most participants recognized condoms as an effective preventive strategy. Similarly, 121 respondents (80.7%) reported that avoiding multiple sexual partners could reduce HIV risk, reflecting good awareness of behavioural prevention measures.

Despite this awareness, practical self-efficacy was comparatively lower. Only 92 respondents (61.3%) reported confidence in their ability to negotiate safer behaviours, including insisting on condom use, refusing unsafe sexual advances, or discussing HIV prevention with a partner. This suggests that a considerable proportion of respondents may lack the communication skills or confidence required to translate knowledge into protective action.

Furthermore, only 79 respondents (52.7%) reported consistently avoiding behaviours perceived as high-risk, indicating that nearly half of the respondents may still engage in or feel vulnerable to risky situations. This finding highlights an important gap between awareness and behavioural implementation.

With respect to health-seeking practices, 98 respondents (65.3%) reported willingness to seek professional counselling or healthcare support when faced with HIV-related concerns. In addition, 96 respondents (64.0%) indicated that they would seek medical advice promptly after potential exposure to HIV.

Preventive knowledge related to maternal transmission remained relatively limited. Only 88 respondents (58.7%) correctly identified all routes of mother-to-child transmission, suggesting weaker practical understanding of specific preventive interventions in reproductive health contexts.

Overall, the findings suggest that while respondents possessed moderate awareness of HIV prevention strategies, confidence in applying such knowledge in real-life situations remained suboptimal. This indicates that knowledge alone may not be sufficient to ensure effective protective behaviour, emphasizing the need for interventions that strengthen practical skills and self-efficacy.

**Table 5.** Preventive Practices and Self-Efficacy.

| Practice | Frequency (%) |
| --- | --- |
| Condom use reduces HIV risk | 112 (74.7) |
| Avoiding multiple partners reduces risk | 121 (80.7) |
| Confidence negotiating safer behaviour | 92 (61.3) |
| Avoidance of risky behaviour | 79 (52.7) |
| Willing to seek counselling | 98 (65.3) |
| Would seek medical care after exposure | 96 (64.0) |
| Correct MTCT preventive knowledge | 88 (58.7) |

### Sources of HIV Information

Respondents reported multiple channels through which they accessed HIV-related information, reflecting the diverse communication platforms available to adolescents.

Schools emerged as the predominant source of HIV-related information, reported by 119 respondents (79.3%). This finding highlights the important role of school-based health education programmes, particularly initiatives such as Family Life and HIV Education (FLHE), in improving HIV awareness among adolescents.

The second most frequently reported source was social media, identified by 98 respondents (65.3%). This suggests that digital platforms have become a major channel for health information dissemination among young people. While social media may improve access to information, it also presents potential risks related to misinformation.

Only 75 respondents (50.0%) reported receiving HIV-related information from parents or guardians, indicating comparatively limited family-based communication regarding sexual and reproductive health matters. This suggests that discussions about HIV may remain sensitive or infrequent in many households.

Traditional mass media also contributed to awareness. Television and radio were reported by 69 respondents (46.0%), showing that conventional media still plays a moderate role in health communication.

Healthcare professionals were among the least reported information sources, with only 54 respondents (36.0%) identifying health workers as a source of HIV-related information. This may indicate limited adolescent engagement with formal healthcare counselling services.

Additional information sources included friends or peers (63; 42.0%) and internet websites/blogs (58; 38.7%), demonstrating the influence of peer networks and online information-seeking behaviour among adolescents.

Overall, the findings indicate that formal educational settings and digital media were the dominant channels for HIV-related information, while parental communication and healthcare-based education played comparatively smaller roles.

**Table 6.** Sources of HIV Information.

| Source | Frequency (%) |
| --- | --- |
| School | 119 (79.3) |
| Social media | 98 (65.3) |
| Parents/Guardians | 75 (50.0) |
| Television/Radio | 69 (46.0) |
| Friends/Peers | 63 (42.0) |
| Internet websites/blogs | 58 (38.7) |
| Health workers | 54 (36.0) |

## Discussion

This study assessed HIV-related knowledge, misconceptions, attitudes, preventive practices, self-efficacy, and sources of HIV information among female adolescents attending a public secondary school in southeastern Nigeria. Overall, the findings demonstrated generally adequate HIV knowledge and moderately positive attitudes toward HIV prevention and testing. However, important misconceptions regarding HIV transmission persisted, while preventive practices and behavioural self-efficacy remained weaker than knowledge and attitudes. Schools and digital media emerged as the dominant sources of HIV-related information.

These findings emphasize the multidimensional nature of HIV prevention among adolescents, where awareness alone may not necessarily translate into protective behavioural practices. Evidence from Nigeria similarly shows that although HIV knowledge and awareness among adolescents have improved, gaps remain in comprehensive understanding, stigma reduction, and testing uptake (Badru et al., 2020; Badejo et al., 2024; Adekunjo et al., 2021). This suggests that HIV prevention interventions should extend beyond awareness creation to include stigma reduction, behavioural skill development, and improved access to adolescent-friendly health services.

### HIV Knowledge Among Respondents

The present study found that respondents demonstrated generally adequate HIV knowledge, particularly regarding sexual and blood-related transmission routes. This suggests that existing HIV awareness campaigns and school-based sexuality education programmes may be contributing positively to adolescent understanding of HIV transmission and prevention.

This finding is consistent with recent Nigerian and regional evidence reporting moderate to high HIV knowledge among adolescents. For example, Badru et al. (2020) found that awareness of major HIV transmission routes was relatively high among young adolescents, although comprehensive knowledge remained suboptimal. Similarly, Ezelote et al. (2024) reported significant improvement in HIV knowledge among in-school adolescents following peer-led health education interventions in Imo State, highlighting the effectiveness of structured school-based educational programmes.

The relatively high knowledge observed in this study may also reflect increased exposure to structured sexuality education within schools. The Federal Ministry of Education Nigeria and NERDC (2003) emphasize HIV and family life education as part of the national curriculum, which may contribute to improved awareness levels. In addition, global evidence suggests that adolescent HIV literacy is improving in many settings, although depth of knowledge remains uneven (UNAIDS, 2024; WHO, 2024).

Despite generally adequate knowledge, weaker understanding of mother-to-child transmission was observed. This suggests that HIV education may emphasize sexual transmission more than reproductive and clinical transmission pathways. Such gaps remain important because incomplete understanding of transmission routes may limit comprehensive prevention literacy among adolescent girls.

### Misconceptions Regarding HIV Transmission

Despite adequate general HIV knowledge, misconceptions regarding HIV transmission remained evident among respondents. Misconceptions relating to casual contact, mosquito bites, and the belief that HIV affects only specific groups persisted.

This pattern aligns with evidence showing that misconceptions remain common among adolescents despite improved awareness. Badru et al. (2020) reported that inaccurate beliefs about HIV transmission were still present among young adolescents in Nigeria and were associated with stigma and poor risk perception. Similarly, Abate et al. (2020) found that incomplete HIV knowledge among young women often coexists with persistent misconceptions, particularly in low-resource settings.

The coexistence of accurate knowledge and false beliefs suggests that information exposure alone is insufficient to eliminate deeply rooted myths. Cultural beliefs, peer influence, and misinformation from informal sources may reinforce misconceptions. Persistent myths surrounding casual contact are especially concerning because they contribute to stigma and discrimination against people living with HIV (Folayan et al., 2022; Adekunjo et al., 2021).

Furthermore, the belief that HIV affects only specific “high-risk” groups may reduce adolescents’ perceived susceptibility. When young people underestimate their vulnerability, they may be less likely to adopt preventive behaviours, increasing infection risk.

### Attitudes Toward HIV Prevention and Testing

Respondents generally demonstrated moderately positive attitudes toward HIV prevention and testing, with most recognizing the importance of HIV screening and preventive health behaviours. This aligns with evidence showing gradual improvement in HIV testing awareness among adolescents and young people in Nigeria, although uptake remains limited by stigma and structural barriers (Badejo et al., 2024; Adekunjo et al., 2021). Akande et al. (2024) further highlight that m-health and digital interventions can improve adolescents’ engagement with sexual and reproductive health services, including HIV prevention messaging.

Despite positive attitudes, stigma-related concerns remained evident. Fear of discrimination and discomfort toward people living with HIV suggest that HIV-related stigma continues to be a major barrier to testing and disclosure. Similar findings have been widely documented, where stigma significantly reduces adolescents’ willingness to access HIV services (Folayan et al., 2022; Adekunjo et al., 2021).

### Preventive Practices and Self-Efficacy

A major finding of this study was the gap between HIV knowledge and preventive behaviour. Although respondents demonstrated reasonable knowledge of preventive strategies, their actual preventive practices and self-efficacy were comparatively weaker.

This knowledge–behaviour gap is consistent with broader evidence from Nigeria and sub-Saharan Africa. Badejo et al. (2024) reported that barriers such as stigma, gender dynamics, and limited-service access significantly influence sexual behaviour and HIV testing among adolescents and young adults. Similarly, Folayan et al. (2022) emphasized that structural and psychosocial barriers often prevent adolescents from translating knowledge into practice.

This suggests that behavioural outcomes are influenced by more than knowledge alone. Peer pressure, emotional factors, social norms, and limited negotiation skills may reduce adolescents’ ability to adopt protective behaviours. Strengthening self-efficacy is therefore essential for improving behavioural outcomes.

### Sources of HIV Information

Schools emerged as the predominant source of HIV-related information, followed by social media, indicating that formal education systems and digital platforms play central roles in adolescent health communication.

This supports evidence that school-based HIV education remains one of the most effective strategies for improving adolescent HIV knowledge (Ezelote et al., 2024). The integration of HIV and family life education into school curricula further strengthens this role (Federal Ministry of Education Nigeria & NERDC, 2003).

The growing influence of digital media is also consistent with recent developments in adolescent health communication, where digital platforms increasingly shape awareness and perceptions (Akande et al., 2024; UNAIDS, 2024). However, these platforms may also expose adolescents to misinformation, potentially contributing to persistent misconceptions.

Parental communication and healthcare workers were less frequently reported as information sources. This may reflect cultural discomfort around sexuality discussions within families and limited adolescent-friendly health service engagement. Strengthening multi-channel communication including schools, families, healthcare providers, and digital platforms is therefore essential for comprehensive HIV education.

### Strengths and Limitations

This study focused on female adolescents, a population particularly vulnerable to HIV infection, and provided a comprehensive assessment of HIV knowledge, misconceptions, attitudes, preventive practices, and information sources. The use of a structured, pilot-tested questionnaire and the school-based setting enhanced data reliability and standardized data collection. However, the study has limitations. Being conducted in a single public secondary school limits generalizability, while the cross-sectional design prevents causal inference. In addition, self-reported responses and convenience sampling may have introduced recall, social desirability, and selection biases.

## Conclusion

This study found that female adolescents demonstrated generally adequate HIV knowledge and moderately positive attitudes toward HIV prevention and testing. However, misconceptions regarding HIV transmission persisted, and preventive practices remained weaker than knowledge and attitudes, highlighting a gap between awareness and behaviour. Schools and social media were the major sources of HIV-related information. These findings suggest the need for HIV prevention interventions that go beyond awareness creation to emphasize comprehensive sexuality education, stigma reduction, and behavioural skill development among adolescent girls.

## Abbreviations

AIDS: Acquired Immunodeficiency Syndrome
ART: Antiretroviral Therapy
FLHE: Family Life and HIV Education HIV: Human Immunodeficiency Virus
MTCT: Mother-to-Child Transmission
PLWH: People Living with HIV
SDG: Sustainable Development Goal
UNAIDS: Joint United Nations Programme on HIV/AIDS
WHO: World Health Organization

## Declarations

### Ethics approval and consent to participate

Ethical approval for this study was obtained from the University of Nigeria Teaching Hospital, Research and Ethics Committee, Ituku-Ozalla, Nigeria (Approval No.: [Insert Approval Number]). Permission to conduct the study was also obtained from the management of the selected secondary school. Written informed consent was obtained from all participants prior to enrolment after the purpose of the study had been clearly explained. For participants below 18 years of age, written informed consent was obtained from parents or legal guardians, while assent was obtained from the participants. Participants were informed of their right to withdraw from the study at any time without any consequences. Confidentiality was maintained by using identification codes instead of names, and all collected data were handled securely and confidentially. The study was conducted in accordance with the ethical principles of the Declaration of Helsinki.

## Consent for publication

Not applicable.

## Availability of data and materials

The datasets generated and/or analysed during the current study are available from the corresponding author upon reasonable request. The data are not publicly available because they contain information that could compromise participant confidentiality.

## Authors’ contributions

RE conceived and designed the study, coordinated data collection, KF performed data analysis, and AO drafted the manuscript. SO contributed to study design, data collection, and manuscript preparation. AO and RE contributed to data analysis and critical revision of the manuscript. All authors critically reviewed the manuscript, approved the final version, and agreed to be accountable for all aspects of the work.

## Acknowledgements

The authors sincerely thank all the students who participated in this study for their valuable contributions. The authors also acknowledge the management, teachers, and staff of the selected secondary school for their cooperation and support during data collection.

## References

Abate, B. B., Kassie, A. M., Reta, M. A., Ice, G. H., & Haile, Z. T. (2020). Residence and young women’s comprehensive HIV knowledge in Ethiopia. BMC public health, 20(1), 1603. 10.1186/s12889-020-09687-1

Adekunjo, F. O., Rasiah, R., Dahlui, M., & Ng, C. W. (2021). The effects of HIV-related stigma on HIV counselling and testing in Nigeria: A mediation analysis. Journal of Asian and African Studies, 56(6), 1196–1211. 10.1177/0021909620960150

Akande, O. W., Muzigaba, M., Igumbor, E. U., Elimian, K., Bolarinwa, O. A., Musa, O. I., & Akande, T. M. (2024). The effectiveness of an m-Health intervention on the sexual and reproductive health of in-school adolescents: a cluster randomized controlled trial in Nigeria. Reproductive health, 21(1), 6. 10.1186/s12978-023-01735-4

Badejo, O., Wouters, E., Van Belle, S., Buve, A., Smekens, T., Jwanle, P., Laga, M., & Nöstlinger, C. (2024). Latent class analysis of barriers to HIV testing services and associations with sexual behaviour and HIV status among adolescents and young adults in Nigeria. PLOS ONE, 19(4), e0300220. 10.1371/journal.pone.0300220

Badru, T., Mwaisaka, J., Khamofu, H., Agbakwuru, C., Adedokun, O., Pandey, S. R., et al. (2020). HIV comprehensive knowledge and prevalence among young adolescents in Nigeria: Evidence from Akwa Ibom AIDS indicator survey, 2017. BMC Public Health, 20, 45. 10.1186/s12889-019-7890-y

Declaration of Helsinki. (2013). World Medical Association Declaration of Helsinki: Ethical principles for medical research involving human subjects. JAMA, 310(20), 2191–2194. 10.1001/jama.2013.281053

Dzinamarira, T., & Moyo, E. (2024). Adolescents and young people in sub-Saharan Africa: overcoming challenges and seizing opportunities to achieve HIV epidemic control. Frontiers in public health, 12, 1321068. 10.3389/fpubh.2024.1321068

Ezelote, C.J., Osuoji, N.J., Mbachu, A.J., et al. Effect of peer health education intervention on HIV/AIDS knowledge amongst in-school adolescents in secondary schools in Imo State, Nigeria. BMC Public Health 24, 1029 (2024). 10.1186/s12889-024-18536-4

Federal Ministry of Education Nigeria, & National Educational Research and Development Council. (2003). Family life and HIV education (FLHE) curriculum for junior secondary schools in Nigeria. Abuja, Nigeria: Author.

Folasayo, A. T., Oluwasegun, A. J., Samsudin, S., Saudi, S. N. S., Osman, M., & Hamat, R. A. (2017). Assessing the knowledge level, attitudes, risky behaviors and preventive practices on sexually transmitted diseases among university students as future healthcare providers in the Central Zone of Malaysia. International Journal of Environmental Research and Public Health, 14(2), 159. 10.3390/ijerph14020159

Folayan, M. O., Sam-Agudu, N. A., & Harrison, A. (2022). Exploring the why: risk factors for HIV and barriers to sexual and reproductive health service access among adolescents in Nigeria. BMC Health Services Research, 22, 1198. 10.1186/s12913-022-08551-9

Gillette, E., Nyandiko, W., Chory, A., Scanlon, M., Aluoch, J., Choudhury, N., Lagat, D., Ashimosi, C., Biegon, W., Munyoro, D., Lidweye, J., Nyagaya, J., Wilets, I., DeLong, A., Kantor, R., Vreeman, R., & Naanyu, V. (2023). Ethical Considerations for Engaging Children and Adolescents Living with HIV in Research in African Countries: A Systematic Review. Journal of empirical research on human research ethics : JERHRE, 18(5), 346–362. 10.1177/15562646231208991

Joint United Nations Programme on HIV/AIDS (UNAIDS). (2024). The urgency of now: AIDS at a crossroads—Global AIDS update 2024. Geneva, Switzerland: UNAIDS.

Joint United Nations Programme on HIV/AIDS (UNAIDS). (2024). The urgency of now: AIDS at a crossroads—Global AIDS update 2024. Geneva, Switzerland: UNAIDS. UNAIDS report page

Launiala, A. (2009). How much can a KAP survey tell us about people’s knowledge, attitudes and practices? Some observations from medical anthropology research on malaria in pregnancy in Malawi. Anthropology Matters, 11(1). 10.22582/am.v11i1.31

Murewanhema, G., Musuka, G., Moyo, P., Moyo, E., & Dzinamarira, T. (2022). HIV and adolescent girls and young women in sub-Saharan Africa: A call for expedited action to reduce new infections. IJID regions, 5, 30–32. 10.1016/j.ijregi.2022.08.009

National Agency for the Control of AIDS (NACA). (2019). Nigeria HIV/AIDS indicator and impact survey (NAIIS) 2018: Technical report. Abuja, Nigeria: National Agency for the Control of AIDS.

Setia, M. S. (2016). Methodology series module 3: Cross-sectional studies. Indian Journal of Dermatology, 61(3), 261–264. 10.4103/0019-5154.182410

von Elm, E., Altman, D. G., Egger, M., Pocock, S. J., Gøtzsche, P. C., & Vandenbroucke, J. P. (2007). The Strengthening the Reporting of Observational Studies in Epidemiology (STROBE) statement: Guidelines for reporting observational studies. PLoS Medicine, 4(10), e296. 10.1371/journal.pmed.0040296

World Health Organization. (2024). Adolescent health. Geneva, Switzerland: Author.

